# ClinGen Variant Curation Interface Workshops: Training Variant Scientists on an International Platform

**DOI:** 10.1101/2025.08.27.25334615

**Authors:** D.I. Ritter, M. Mandell, C Preston, M DiStefano, SM Bryant, N Carstens, RS Julius, A Lumaka, M Hedge, J Ngeow, J Morales, M.W. Wright, H.L. Rehm, S.E Plon

**Affiliations:** Baylor College of Medicine; Stanford University; Medical and Population Genetics, Broad Institute of MIT and Harvard, Cambridge, MA USA; University of the Witwatersrand/NHLS Division of Human Genetics; University of Cape Town; University of Kinshasa; Revvity Inc; Lee Kong Chian School of Medicine, Nanyang Technological University Singapore and Cancer Genetics Service, National Cancer Centre Singapore; National Human Genome Research Institute; Center for Genomic Medicine, Massachusetts General Hospital, Boston, MA, USA

**Author notes:** Corresponding Author: Dr. Deborah Ritter.

## Abstract

**Introduction:** The Clinical Genome Resource (ClinGen) is creating a central resource of clinically relevant genetic knowledge to improve genomic medicine. Dissemination and use of the ClinGen Resource is essential to ensure broad community uptake. We report on experiences and sustained use of ClinGen tools through engaging international genetics groups based in India, Africa and Singapore in variant classification training workshops using the ClinGen Variant Curation Interface (VCI).

**Methods:** We developed pre and post workshop questionnaires and analyzed ClinGen tool use following the workshops. We evaluated organizational aspects and costs of creating a dedicated ClinGen VCI instance for each workshop.

**Results:** The workshops yielded >200 participants, with local scientists as essential participants. While ∼55% of participants were unfamiliar with variant classification, we found ∼79% were likely to use the VCI after the workshop. Further, we identified about ∼10% of workshop participants created permanent accounts. We estimate costs at ∼$3 per VCI instance.

**Discussion:** Our efforts highlight the yield of international workshops to sustained use of ClinGen’s curation tools, and identify areas for future consideration such as creating user-groups by experience level, and the importance of local scientist engagement in workshop deployment and organizational aspects.

## Introduction

Training and outreach to the international genetics community can foster relationships that encourage data sharing and cooperative partnerships for the improvement of genomics knowledgebases. The Clinical Genome Resource (ClinGen) is an NIH-funded consortium that provides an authoritative database and resource for clinically relevant genomic information^1^. ClinGen frequently offers educational outreach efforts to engage users from the scientific community, which increases usage and feedback that can, in turn, improve offerings^2^. Here, we report on three workshops developed and delivered to over 200 researchers and participants from international genetics groups in India, Africa and Singapore. Each of these genetics groups have significant numbers of participants from low-to-middle income countries (LMIC) with nascent genetics networks and are actively seeking enhanced local training opportunities for advanced clinical genomics ^3^. During the recently held first meeting of the Ugandan Society for Human Genetics and Bioinformatics and the 22nd Congress of the African Society for Human Genetics, ClinGen and Human Hereditary and Health Africa (H3Africa) developed virtual genomics training to supplement the meeting program. Related efforts through ClinGen and Human Hereditary and Health Africa have developed additional virtual genomics trainings available on the ClinGen website. Similarly, the Society for the Indian Academy of Medical Genetics has held annual conferences and invites scientists from the USA to enhance genetic training networks and opportunities. Likewise, the SingHealth Duke-National University of Singapore National Cancer Center, held a conference with associated workshops featuring USA-based scientists to encourage data sharing and further develop local cancer genomics analysis skills. To increase global consistency of variant classification through encouraging broad use of the ClinGen Variant Curation Interface and other tools and products in the international genetics community, we collaboratively designed three workshops with the international hosts, using iterative improvements from participant feedback, and resulting in modular and reusable offerings that enhance local training capacity ^4, 5^. We describe our workshop design and how participant feedback across the three training groups was incorporated to improve subsequent training developments, as well as summarize participant responses and outcomes.

## Materials and Methods

### Participant Numbers

Participation was tracked through workshop registrations and the Variant Curation Interface accounts. The Society for the Indian Academy of Medical Genetics Conference (hereafter, SIAMGCON) had 83 participants, Human Hereditary and Health in Africa (H3Africa) 74 and Duke-NUS NCCS (hereafter, NCCS) 44, for a total of 201. SIAMGCON had no formal pre/post survey, though informal and spontaneous feedback informed development of the next two workshops, registrations and pre/post questionnaires.

### Questionnaires

Registrations for H3Africa and NCCS included 9 pre-workshop questions on the participant’s educational background and baseline familiarity with variant curation. Post-workshop questionnaires of 13-18 questions were sent to H3Africa and NCCS, with a 46% and 32% response rate, respectively. An additional post-workshop questionnaire (referred to as “External NCCS Questionnaire”) was circulated by the meeting organizers and received 50 responses. For “Participant Satisfaction” the responses from the External Questionnaire were harmonized and combined with those of the workshop-specific questionnaire due to the higher response rate.

### Mapping and Harmonizing

Each workshop was unique in location, participants and leading members, and questionnaires had slight differences (i.e. responses in Likert scale vs categories) requiring minor content mapping to merge, described here. *Participant Country of Residence:* Data from H3Africa registration “current country of residence” was combined with the NCCS Workshop Pre-Test “country of your primary institution”. *Participant level of familiarity with variant classification:* H3Africa data from the pretest “Participant level of familiarity with performing variant classification” and the NCCS pretest “Have you used the Standards and Guidelines for the Interpretation of Sequence Variants from The ACCMG?” was combined by mapping terms as follows: From the H3Africa post-survey, “unfamiliar” and “this was my first time” mapped to “unfamiliar.” For the NCCS pretest we used the following: “very frequently” and “frequently” mapped to” very familiar”, “moderately” mapped to “familiar” and “infrequently” or “never” mapped to “unfamiliar”. *Likelihood of Using the VCI post-workshop*: Data from the H3Africa post-survey “Will you use the ClinGen Variant Curation Interface more often after this workshop?” was combined with the NCCS Post Survey “After the workshop, how likely are you to use the ClinGen Variant Curation Interface?”. For NCCS, responses marked as 4 or a 3 were mapped to “likely”, and 2 or 1 mapped to “not likely.” *Interest in ClinGen Involvement:* NCCS responses were mapped to those of H3Africa using the following: “I am interested but now right now,” “I hope to join an expert panel” or “I plan to volunteer as a biocurator” were mapped to “yes”, and responses of “not sure yet” were mapped to “maybe.” *Participant overall workshop satisfaction:* Data from H3Africa post questionnaire “Overall, how satisfied were you with the content presented in this workshop” and NCCS External questionnaire “Do you feel the ClinGen hands-on workshop was well planned?” NCCS responses of “strongly agree” and “agree” were mapped to “very satisfied” and “somewhat satisfied.”

### VCI Platform Considerations

For each workshop, several instances of the VCI were created on the Amazon Web Services (AWS) cloud platform. Instances are deployed using AWS CloudFormation templates, an AWS Infrastructure as Code (IaC) service. VCI instances were maintained for 4 months after the workshop to ensure users had time to review curated data. The overall cost to create and maintain 10 VCI instances for 4 months is estimated to be about $30.00 total ($3.00 per instance) in space/processing and one week of FTE effort; deployments, loading of workshop users into their respective instances, and deletion of the instances. *Registration and Affiliations:* Workshop registrants were asked to provide an email, first and last name as well as an institution. Accounts were created for participants in advance and assigned to “affiliations”, which are sub-groups of 5-10 workshop attendees allow members of an affiliation to co-work on a single variant curation within the VCI platform. A few extra affiliations and generic accounts were generated, to accommodate late registrations or for technical issues with email/logins. *VCI Technical Assistance:* For all three workshops, the VCI developer team reserved a 1-hour session in advance of the workshop as a virtual drop-in technical session to receive login help or answer questions about using the VCI. At least one member of the team was available on-call virtually during the workshops. *VCI Production accounts and affiliations:* To assess if VCI accounts and affiliations were established in the production version, we queried the registrant emails against the production database, and institutions. Additionally, we manually reviewed non-ClinGen affiliations established in the VCI Production Affiliations.

## Results

### Workshops Overview

#### SIAMGCON

The first workshop was a half day workshop developed and conducted with the Society for the Indian Academy of Medical Genetics Conference (SIAMGCON). The connection between ClinGen and SIAMGCON was forged through the hereditary cancer clinical domain (the PTEN Variant Curation Expert Panel). To develop the content, we met with the organizing participants and chose (or curated) variants of interest for the participants, as well as progressively challenging variant examples (TABLE 1). The workshop was designed for in-person participation only, due to internet bandwidth capacity at the center. Attendees completed a short survey to register for the workshop. An instance of the ClinGen Variant Curation Interface was created, and 10 affiliations were generated. An affiliation allows members to curate as a group, and to view progress and data on a variant curated in real-time. The attendees were split into groups of 5-10 participants for each affiliation. Prior to the workshop, each participant created an account in the VCI, and their login information was linked to the affiliation. A few additional affiliations were created to accommodate unregistered participants. The workshop progressed as shown in Figure 1, with variants divided into roughly three sessions of progressively challenging curations. The affiliation groups worked separately on variant curations, returning to the full group to share results and discussion, and proceeding to the next set. This progressively challenging approach helped to balance the range of participant experience, providing introductory variants for inexperienced participants and challenging variants for experienced participants. The workshop received positive feedback from attendees and resulted in the subsequent creation of 11 user accounts in the production VCI. However, at the time we did not collect formal pre/post questionnaires on VCI use.

**Table 1:**
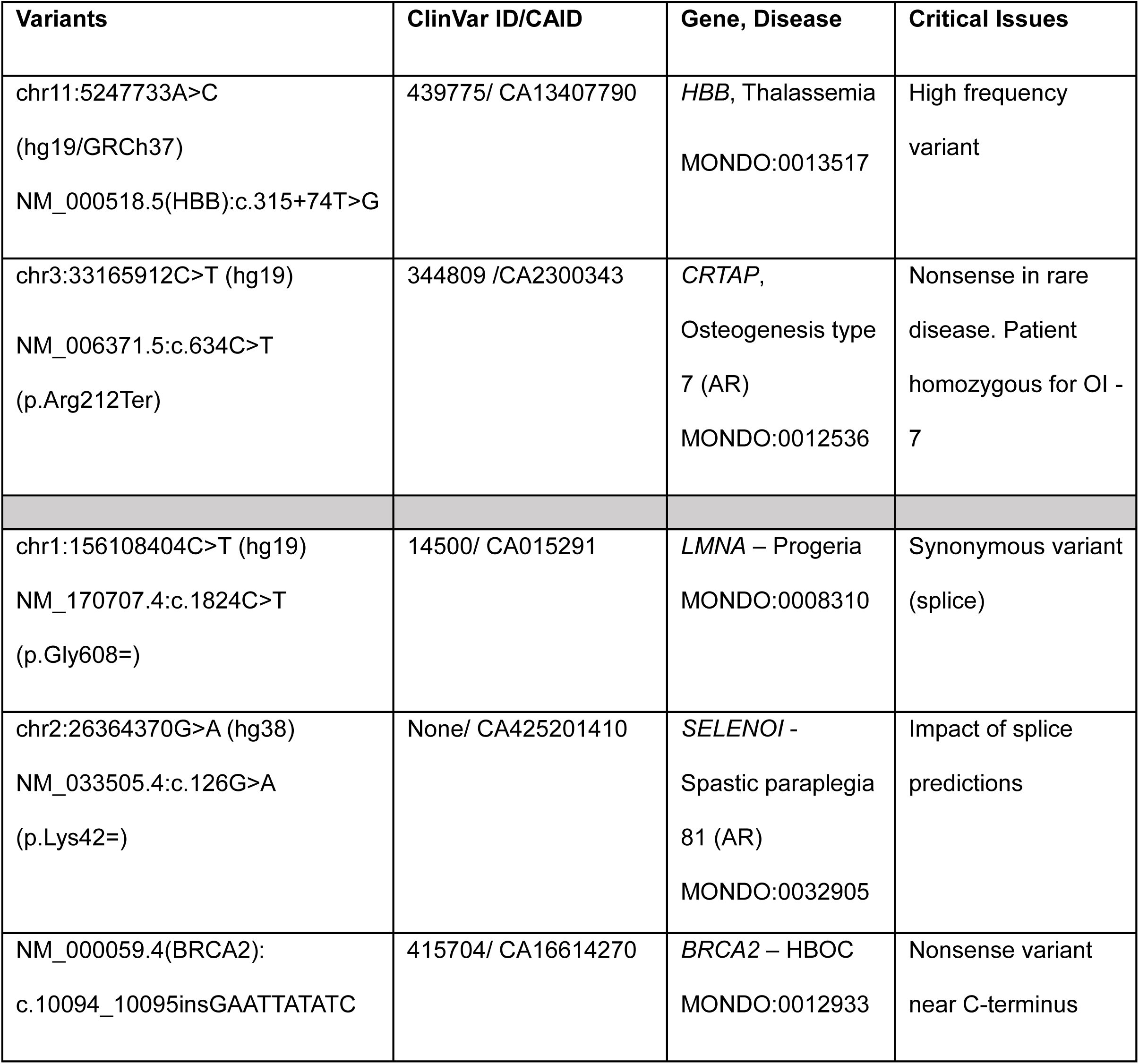
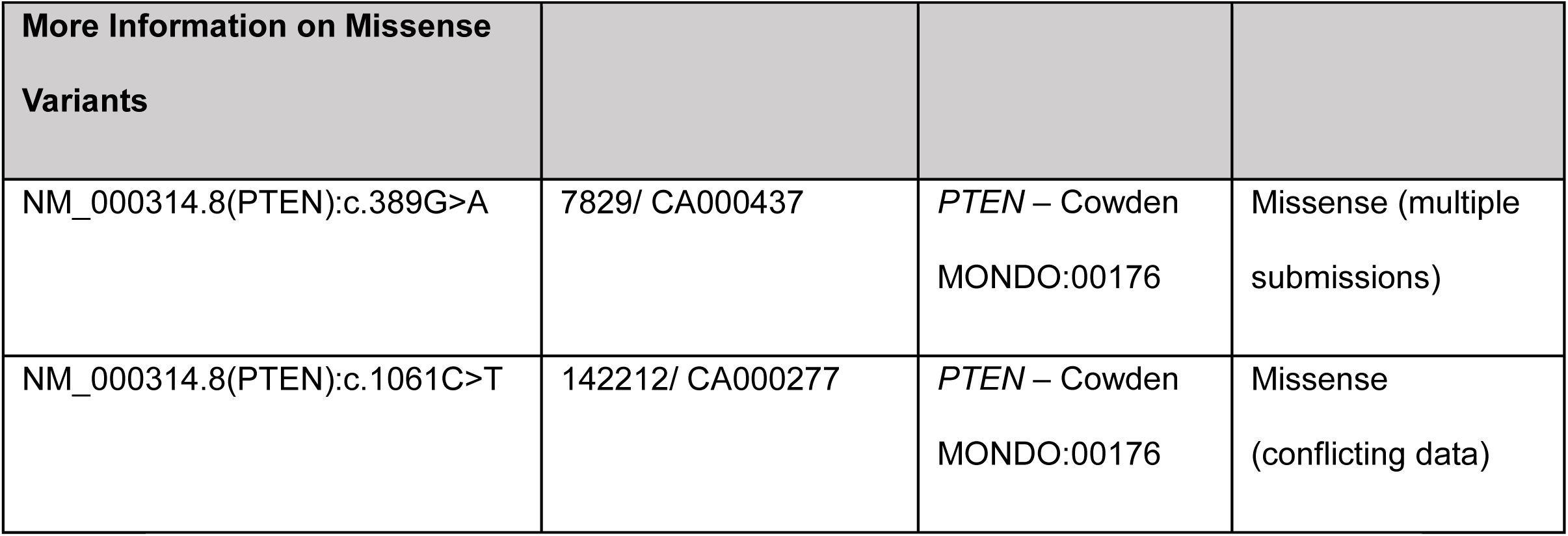
Example Variant Table. Each group leader received a similar variant table with the syndrome, variant type and details about “critical issues”. The variants were ordered from least to most challenging.

**Figure 1:**
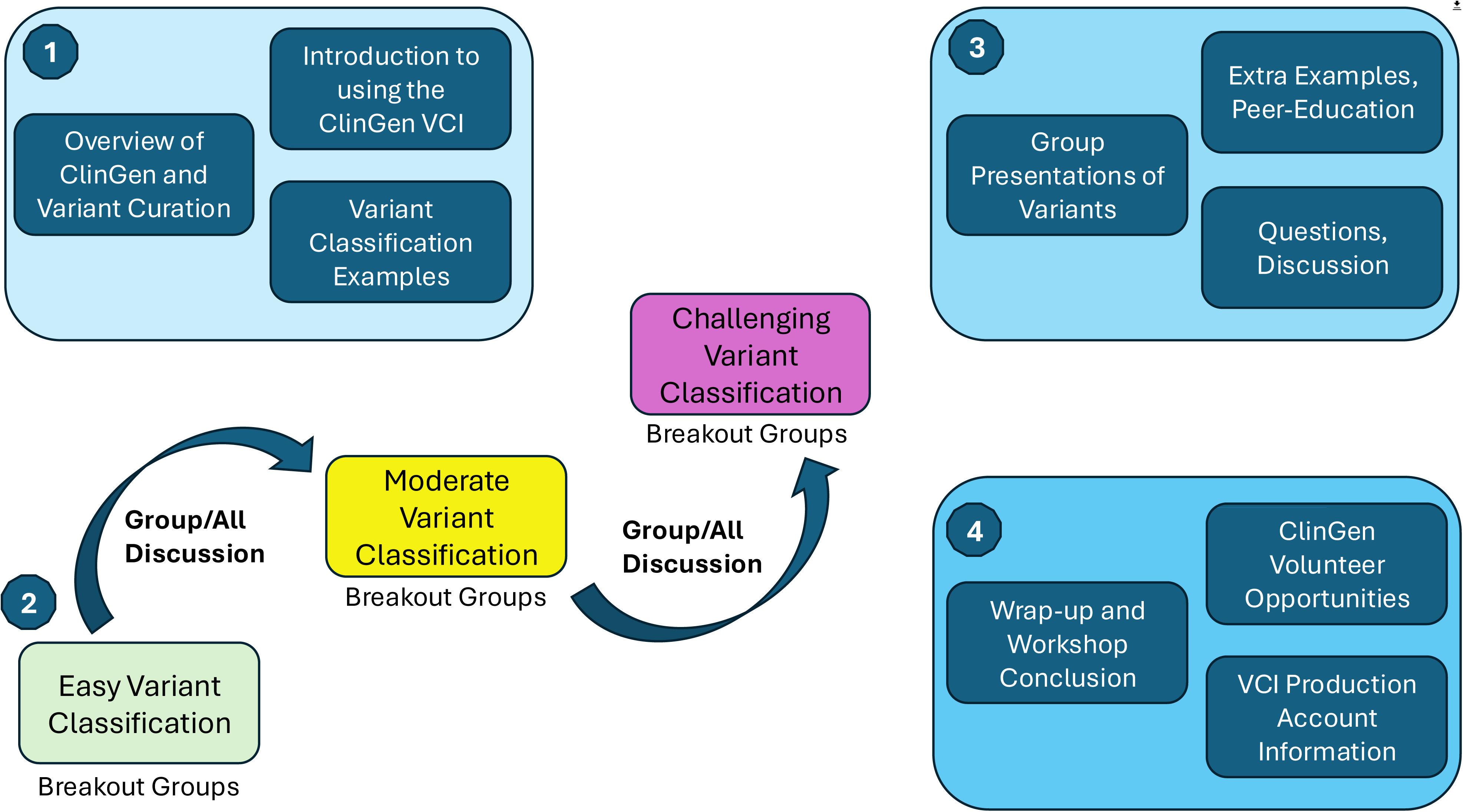
Overview of VCI Workshop Design: (1) Workshops include an overview of variant classification with specific, concrete examples, and a brief introduction to the ClinGen VCI. Following this (2), progressively challenging variants are curated in break-out sessions or small groups in-person, with an experienced team-lead, and all-group discussion between sets. Next (3), small groups present work to the full workshop, with emphasis on peer-education, questions and discussion. Finally (4), the workshop concludes with opportunities to volunteer and information on obtaining accounts in the production version of the VCI.

#### H3Africa

Shortly after the SIAMGCON workshop, we planned a second 3-hour workshop as a virtual offering with H3Africa in the spring of 2023. Through a longstanding partnership and working group with the H3Africa Rare Disease WG, ClinGen has offered several past workshops on genetics topics, including training on use of ClinGen tools and website. These workshops informed our VCI-focused training workshop, as did the prior SIAMGCON workshop. We chose to keep the same progressively challenging approach, as well as small groups in Zoom break-out rooms to focus on curation, with conversational sharing of curated variants between groups. We again built the variant list with input from participants. We developed a pre-workshop questionnaire to assess familiarity with variant curation and ClinGen VCI and tools use, and a post-workshop questionnaire to assess interest in ClinGen participation as well as developing an ongoing affiliation in the VCI. At the SIAMGCON workshop, many participants commented on the benefit of having a “team lead” familiar with the materials and platform in each group. We used this feedback to invite experienced ClinGen biocurators to be team leads for each of the break-out groups in the H3Africa Workshop. All participants were invited to screen share to receive help with curating or to ask questions. After about 15 min time, we returned to the main Zoom room to share results from each team and discuss the curations. For this workshop we again created a custom instance of the VCI. We provided workshop participants the materials and information about how to use the VCI in advance. Our post-workshop questionnaire included questions about use of the VCI and interest to participate in ClinGen or to establish an affiliation in the production version of the VCI.

#### NCCS

The NCCS workshop in Singapore was initiated through a connection between the Hereditary Cancer Clinical Domain chair and an international scholar from Singapore participating in a variant curation Expert Panel. The workshop was in the Fall 2024 and was modeled as a mix of the two prior workshops: developed in conversation with NCCS participants and interests and conducted in-person with virtual support from the VCI team. The workshop was half a day and followed the same model of progressively challenging variants. As discussed in the results, feedback from the H3Africa workshop was put into use for the NCCS workshop, especially in group composition. Again, the progressively challenging variants were informed by participant interests, and each group had an in-person biocuration lead.

### Workshop Pre and Post Survey Responses

In total we had 201 participants across the three workshops (SIAMGCON had 83 participants, H3Africa = 74 and NCCS = 44). We obtained the country of residence from H3Africa and NCCS workshops (Figure 2A and B), which shows participants from at least 13 countries (not including SIAMGCON, which would likely add additional countries). This indicates a broad reach for these two organizations, although more attendees directly in Singapore attended the workshop local to Singapore.

**Figure 2:**
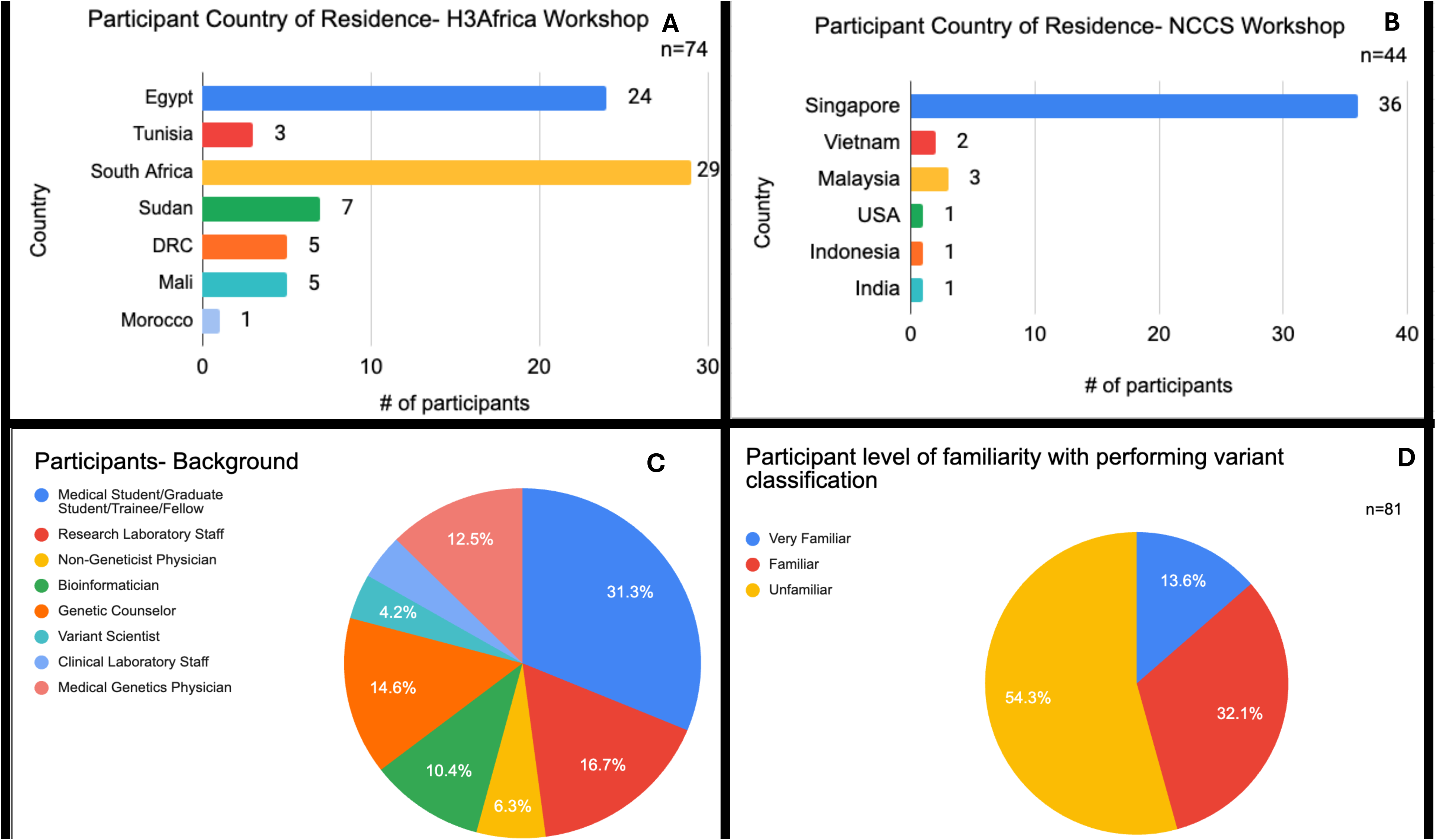
Participant Overview. (A) Country of Residence (H3Africa only), (B) Country of Residence (NCCS only), (C) Educational or Career Background and (D) Self-Reported Familiarity with Variant Classification. Note: Figures C and D combine H3Africa and NCCS data.

Participants came from a variety of backgrounds connected to the genetics profession (Figure 2C), with the largest group (∼30%) reporting as students/trainees/fellows, and the next highest as follows: research laboratory staff (∼17%), genetic counselors (∼15%), medical genetics physicians (13%) and bioinformaticians (10%). Interestingly, clinical laboratory staff or variant scientists made up a very small percent (about 4% each) of the attendees. This indicates that the background of participants is slightly skewed towards having training in genetics but lacking specific variant curation or classification experience; this is very informative to understanding the baseline training in variant classification needed to develop critical analysis skills and use of the Variant Curation Interface in one workshop session, and may support efforts to use pre-workshop materials or testing to assist with background information for the session.

To gauge user familiarity with the ClinGen Variant Curation Interface and curation, we asked about the participant level of familiarity with performing variant classification (Figure 2D). Almost 55% of the attendees from H3Africa and NCCS were unfamiliar with variant classification, with about 30% characterizing their experience as familiar with variant classification and the remaining 13% reporting as “very familiar” with variant classification. This indicates a broad range of user experience across the participants and a need to stratify the learners into similar peer groups during the breakout sessions.

We asked how likely participants were to use the workshop information in their daily career, and around 90% affirmed they were likely or somewhat likely to use it, with 10% neutral and none responding that they would not use the workshop information (Figure 3A). We specifically asked about the likelihood of using the Variant Curation Interface, the platform on which participants performed curations after their introduction to it through the workshops (Figure 3B). Overall, ∼79% stated they would be more likely to use the VCI after the workshop, although we note that the percentage differed between the two groups, with the H3Africa group with 88% confirming likely use and only 57% of the NCCS group confirming likely use. We additionally asked whether participants would consider getting involved in ClinGen by joining a working group or variant curation expert panel, with around 90% replying they are interested to join, and 6% declining with 4% uncertain (Figure 3C). Overall, these responses are well reflected by the number of accounts established in the VCI production version. About 10% (25/236) new accounts were established in the production VCI based on these workshops, with a total of 16% (39/236) accounts when combining newly established and existing accounts from workshop participants. Currently, we are unable to track whether participants in our workshops eventually volunteer in expert panels (through the ClinGen Community Curation efforts).

**Figure 3:**
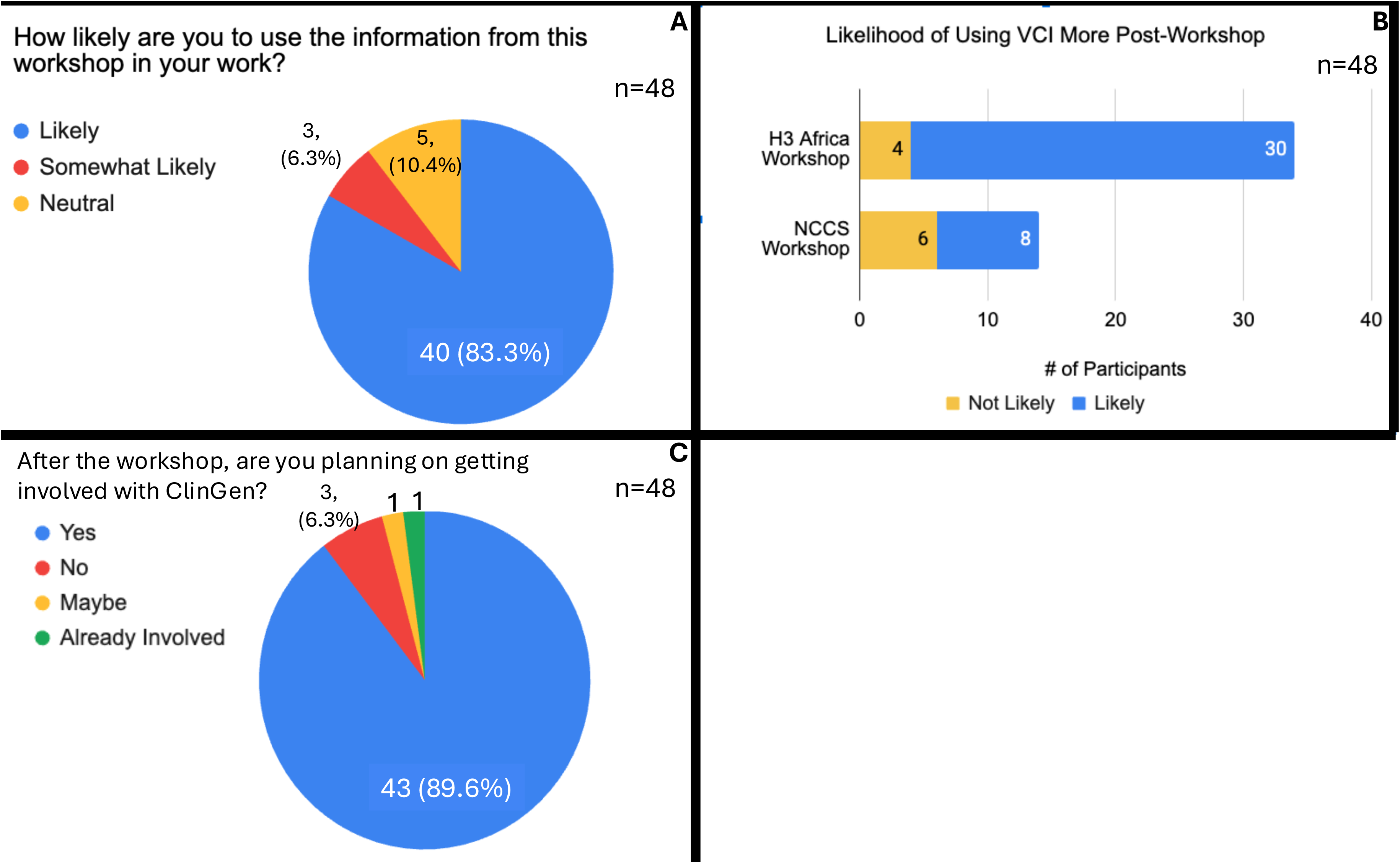
Participant Use of Workshop Content & ClinGen. (A) Use of workshop content in work (B) Likelihood of using the ClinGen VCI post-workshop and (C) Interest in being involved in ClinGen

Participants were asked about whether the workshops provided adequate time, their overall satisfaction with the workshop, and to give free text responses for any other thoughts on the workshops. While 80% agree or were neutral, 20% disagreed and felt more time was needed (Figure 4A). Overall satisfaction with the workshops was high, with 98% being satisfied or somewhat satisfied, 1 respondent being “neutral’ and none reporting they were dissatisfied with the workshops (Figure 4B). About 80% of respondents confirmed they would like an annual workshop on variant curation, with 17% requesting every other year (and 1 no response) (Figure 4C). Finally, free text responses were summarized to identify common trends: 7/13 responses reported that the workshop was too short or felt rushed, as well as 7/13 requested more introductory material – which would help create balance between experienced learners and novice learners (Figure 4D). While the responses were anonymous, we hypothesize that those participants less familiar with variant curation and curation platforms likely needed additional time and training, while those who felt the time was adequate may be more advanced participants. Overall, the results reinforce continued workshop development, with attention needed to clarify the prerequisites required for attendees to maximize workshop learning.

**Figure 4:**
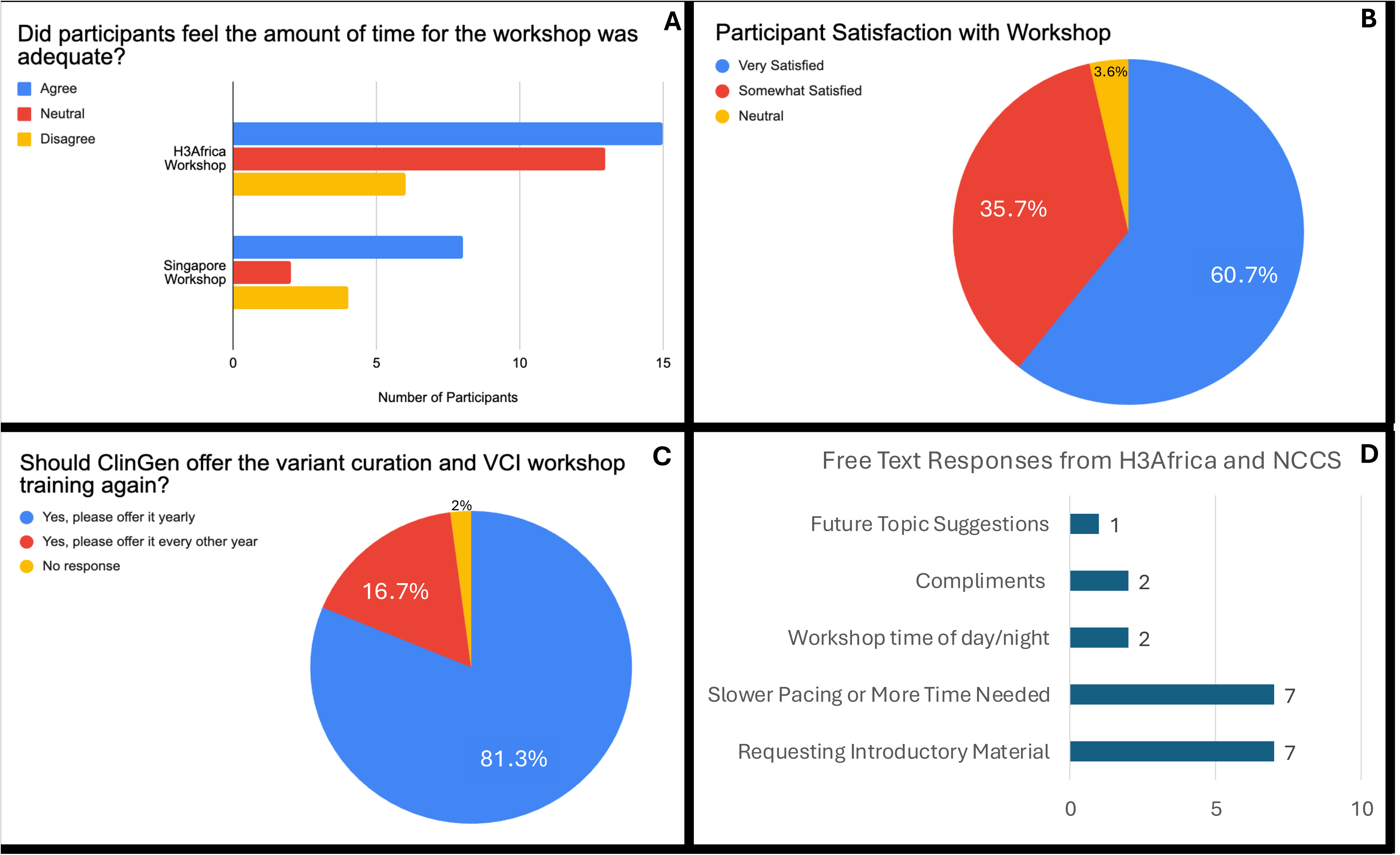
Participant Satisfaction with Workshop. (A) Participant response on workshop length (B) Participant overall satisfaction and (C) Interest in recurrent ClinGen workshops (D) Summarized free-text responses from H3Africa (12) and NCCS (1)

## Discussion

Through three ClinGen variant curation workshops we engaged three international genetics groups with over 200 participants across over 13 countries. The workshop design contained several features that resulted in high levels of participant satisfaction such as follows: progressive variant challenges, small group work, participant-driven variant collection and a supportive curation platform designed for co-working on a variant curation. Feedback from workshops attendees played a critical role in iterative development of subsequent workshops and will continue to inform future workshops. However, we identified key challenges in each workshop, which we review here. A main challenge was encountered when grouping participants by country (or institute) versus by experience. The choice of grouping by country was made per request of the workshop organizing committee mainly for the purpose of keeping participants together by institutes and regionally, to help encourage local networking. However, some of the free text responses reflected incongruity between learners (advanced vs novice) and workshop pacing based on familiarity with content. This feedback guided the next workshop (NCCS), where we created groups based on self-reported familiarity with variant classification. While it is important to consider the requests of organizing participants, we have learned to prioritize user experience when creating workshop teams. For comments related to the pacing and content length, the feedback may relate to experience level or to potential language barriers, as many participants spoke English as a second language. Our questionnaires did not ask about English language fluency, and in subsequent international workshops we will include this question. Ensuring a slower pace and assessing time constraints vs content included will be prioritized in subsequent workshops. For all three workshops multiple other factors were in place when planning workshop content and length, including other competing or parallel workshops (at the SIAMGCON and NCCS workshops), as well as large differences in time zones between participants and presenters (for the H3Africa workshop). This led to challenges in attention and absorbance of materials. From free text feedback, we identified the continued need to provide participants with advance materials on VCI use and login. Each workshop held a technical support session in the day or two prior to the workshop. Several login issues which may have used valuable workshop time were resolved in advance – mostly related to incorrect registration emails or email casing (capital vs lower case).

As noted in the methods section, the creation and 4-month maintenance for a single VCI instance is currently around ∼$3.00 per instance. If the instance is maintained for less time, the cost may further decrease. The administrative overhead relating to instance and account creation was not accounted for completely and has decreased with each deployment of the workshops, although it does require some manual steps in account entry and creation. While we reason that the technical cost may not be a substantial barrier, it is worth considering that many additional elements such as full-time employee (FTE) and expert hours in workshop organization, planning and deployment were not accounted for in the cost above.

Overall, our workshops yielded newly established accounts from ∼10% of participants, with ∼16% of total participants maintaining VCI accounts. These longer-term affiliations and users are a significant outcome, as they signify an ongoing network to consider when developing new workshops. The experience of collaborative design through developing VCI training workshops with international genetics groups has increased the VCI user base while providing valuable feedback for developing VCI training on the new (v4) ACMG/AMP/CAP/ClinGen variant classification guidelines. These workshops were part of ClinGen’s efforts to foster international collaborations in genomic data sharing by engaging researchers and clinicians from diverse regions, with the aim to improve global genomic knowledgebases and standardize variant classification practices across borders. We note that a key factor in the success and sustainability of the effort was the collaboration between a ClinGen and motivated local partners (such as the H3Africa Rare Disease Working Group). This was instrumental in identifying and engaging impactful trainee groups, as local collaborators brought deep contextual knowledge, trusted community relationships, and an understanding of regional challenges^6^. The collaborative model significantly contributed to the successful delivery of the training and laid the groundwork for long-term integration of skills into clinical and research practice. As the regional genomics infrastructure continues to develop, these workshops could evolve into core training components for local research initiatives and healthcare systems. The early evidence of participants applying their knowledge, maintaining engagement, and seeking further collaboration with ClinGen suggests a promising foundation for scaling and sustaining this model in similar low-resource settings.

## CReDIT Author Crediting

1. Conceptualization – DIR, AL, SEP, RSJ
2. Data curation – DIR, SMB, CP, MM, MH, JN, MD
3. Formal analysis – DIR, SMB, CP, MM
4. Funding acquisition HR, SEP, JM, MWW
5. Investigation – DIR, SMB, CP, MM, JN, MD, MH
6. Methodology DIR, SMB, CP, MM, JN, MD, MH
7. Project administration - JM, SEP, HR, MWW, AL, RSJ
8. Resources MWW, MM, CP, MD, MH, JN, DIR
9. Software MM, CP, MWW
10. Supervision AL, RSJ, SEP, HR, MWW, JM
11. Validation – Not applicable
12. Visualization SMB, DIR
13. Writing-original draft DIR
14. Writing-review & editing DIR, NC, SEP, AL, RSJ

## Data Availability

All variants in the study are available online at ClinVar and educational workshop materials are available on the Clinical Genome Resource website or by author request.

https://clinicalgenome.org/

## References

1. ClinGen Consortium Electronic address: splon@bcm edu and ClinGen Consortium. The Clinical Genome Resource (ClinGen): Advancing genomic knowledge through global curation. Genet Med 2025. DOI: 10.1016/j.gim.2024.101228.

2. Bonham VL and Green ED. The genomics workforce must become more diverse: a strategic imperative. Am J Hum Genet 2021. DOI: 10.1016/j.ajhg.2020.12.013.

3. Sharaf A, Nesengani LT, Hayah I, et al. Establishing African genomics and bioinformatics programs through annual regional workshops. Nat Genet 2024. DOI: 10.1038/s41588-024-01807-6.

4. Preston CG, Wright MW, Madhavrao R, et al. ClinGen Variant Curation Interface: a variant classification platform for the application of evidence criteria from ACMG/AMP guidelines. Genome Med 2022. DOI: 10.1186/s13073-021-01004-8.

5. Tekola-Ayele F and Rotimi CN. Translational Genomics in Low- and Middle-Income Countries: Opportunities and Challenges. Public Health Genomics 2015. DOI: 10.1159/000433518.

6. Sirisena ND and Dissanayake VHW. Strategies for Genomic Medicine Education in Low- and Middle-Income Countries. Front Genet 2019. DOI: 10.3389/fgene.2019.00944.

